# The impact of USAID and CDC funding cuts on HIV incidence and mortality in KwaZulu Natal, South Africa

**DOI:** 10.64898/2026.02.18.26346597

**Authors:** Sheela V. Shenoi, Anthony P Moll, Yao-Rui Yeo, Petra Zama, Gavin George, Neo Morojele, John Mbaya, Kay Govender, Henry Sunpath, Sibusiso Gasa, Charmaine De Wet, Mellesia Jeetoo, Themba Ndabandaba, Dyanna Charles, R Scott Braithwaite

**Affiliations:** AIDS Program, Yale School of Medicine, New Haven, CT, USA; Philanjalo NGO, Tugela Ferry, KwaZulu Natal, South Africa; Department of Population Health, New York University, New York, NY, USA; KwaZulu Natal Department of Health, KwaZulu Natal, South Africa; Health Economics and HIV and AIDS Research Division, University of KwaZulu Natal, Durban, South Africa; University of Johannesburg, Pretoria, South Africa; School of Clinical Medicine, University of KwaZulu Natal, Durban, South Africa

**Keywords:** HIV, USAID, CDC, restoring funds, mortality, incidence

## Abstract

**Background:** Abrupt cessation of USAID and CDC resources to KwaZulu Natal province in South Africa, threatens the progress over decades to address HIV.

**Methods:** We used a previously developed validated HIV transmission model with input from the KZN Department of Health and local stakeholders to estimate impact of funding cuts on HIV incidence and mortality at 12-months and through 2030. We applied the model to estimate the impact of restoring funds on HIV incidence and mortality.

**Results:** HIV incidence increased at 12 months and through 2030 by 3.4% and 22.8%, leading to 35,300 and 116,100 additional infections, and 12,800 and 42,300 additional deaths, respectively. Restoring funding after a 12-month pause, reallocated to focus on long-acting PrEP, would avert 12,600 new infections.

**Conclusion:** This model application demonstrates that the sudden cessation of USAID and CDC commitments in the largest HIV epidemic in the world leads to increased incidence and mortality and threatens decades of progress in KZN, South Africa. Restoring funding within 12 months and increasing efficiency of HIV interventions can reestablish KwaZulu Natal province, South Africa’s trajectory toward EHE goals.

## INTRODUCTION

South Africa is home to the one of the world’s largest HIV epidemics. KwaZulu-Natal (KZN) province has the largest antiretroviral program in the country and the greatest proportion of people with HIV (PWH) with viral load (VL) suppression in the country.(1) South Africa, overall and KZN in particular, has benefited tremendously from the United States Agency for International Development (USAID) and the U.S. Centers for Disease Control (CDC) over the last two decades. In South Africa, HIV prevalence has decreased from 16.7% to 12.6% and incidence has declined from 2.7% to 0.44% over the last two decades(2, 3) raising life expectancy from 46 years in 2000 to 62 years in 2023.(4) Over the past five years, the authors have conducted Participatory Policy-guided Modeling (PPM) to inform resource allocation for KZN across a broad suite of HIV treatment and prevention programs by the South African Department of Health and its major external funders. PPM is a structured, implementation-focused adaptation of decision analytic modeling in which decision-makers and other stakeholders frame model questions, select model inputs, and iteratively review model results.(5-8) The funding cuts to CDC and USAID on January 25th, 2025 threatened advances in the HIV epidemic, requiring a rapid reassessment of programmatic vulnerability and potential mitigation strategies. This study seeks to determine the projected impact of USAID and CDC funding cuts on HIV incidence and mortality in KwaZulu-Natal province, South Africa at 12 months and the 5-year interval through 2030, and to what extent restoring or reallocating resources, particularly toward long-acting PrEP, may mitigate these effects.

## METHODS

We previously developed a HIV transmission model from an initial model developed to be applicable across southern African settings,(9-11) and adapted to KZN based on KZN specific inputs (Table 1) and assessing calibration to HIV incidence, prevalence, and HIV-related mortality (Appendix). We obtained input from annual in-person prioritization workshops with KZN Department of Health and local stakeholders, including clinicians, academics, and civil society representatives.(9) Participants underwent structured preference measurement using fixed point allocation to identify the most important strategies for addressing HIV in KZN. Strategies were prioritized based on perceived feasibility, acceptability, importance, and budgetary implication. We applied this previous model to evaluate HIV incidence and mortality if funding were to be paused for 12 months and through 2030. We stratified results by funding cuts involving USAID-versus CDC-implemented interventions (Table 2) using programmatic data and expert opinion gathered during a workshop in Durban, South Africa, on March 6-7^th^, 2025. We considered whether offsets would occur from task-shifting and reallocations of domestic resources.

**Table 1.**
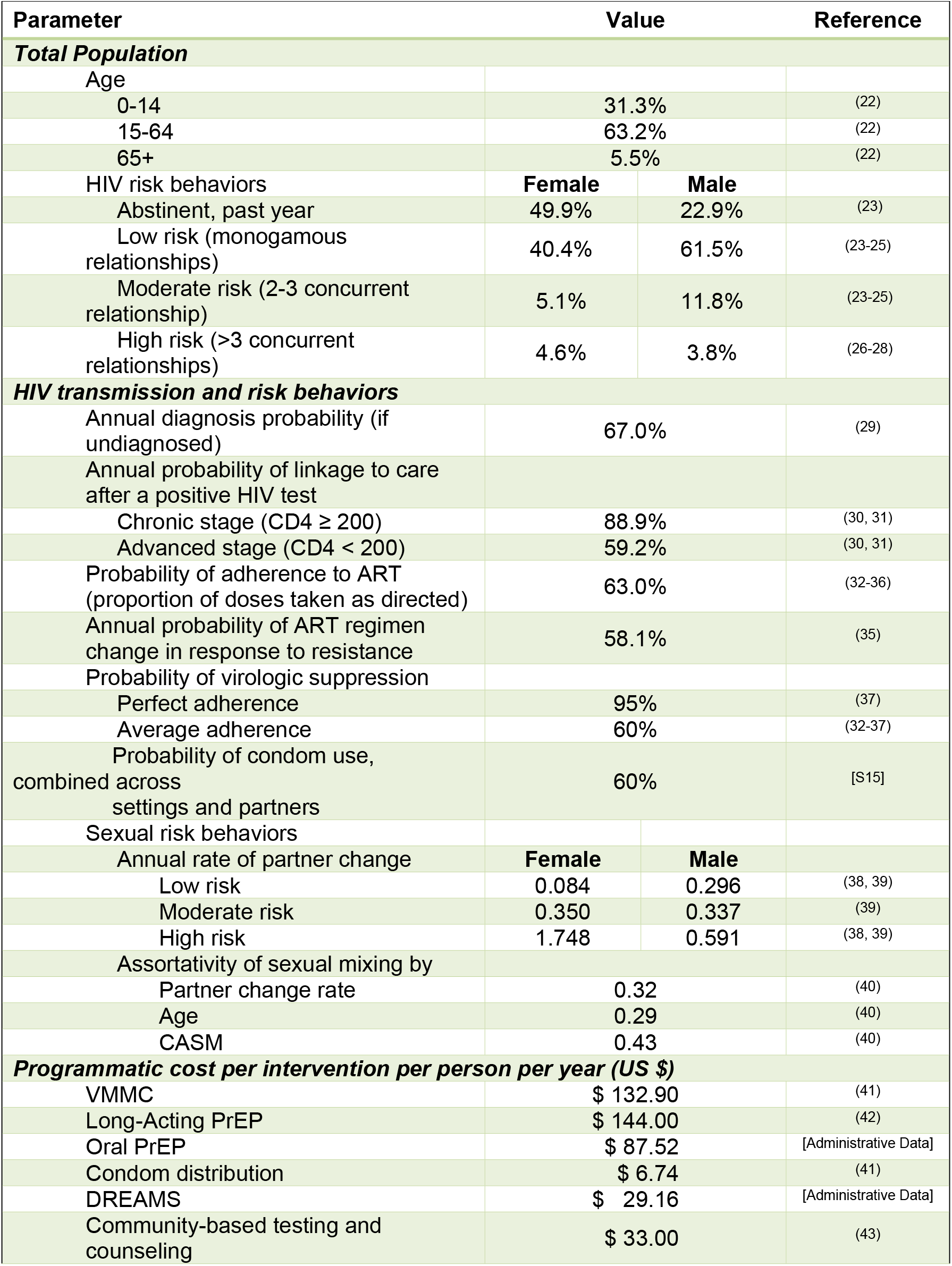
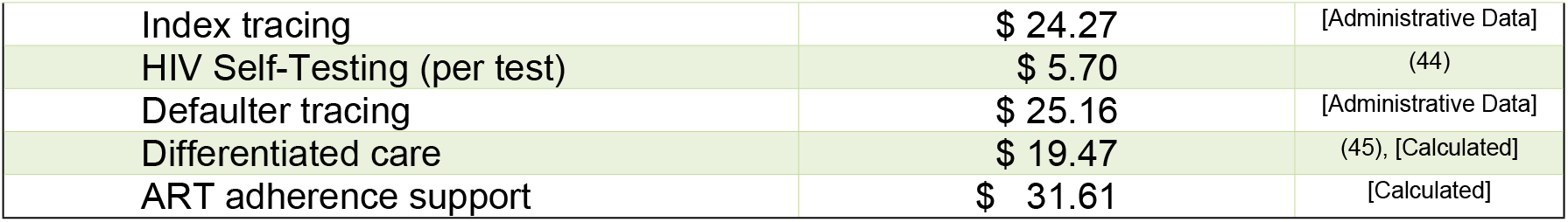
Key Model Inputs.

| Parameter | Value |  | Reference |
| --- | --- | --- | --- |
| **Total Population** |  |  |  |
| Age |  |  |  |
| 0-14 | 31.3% |  | (22) |
| 15-64 | 63.2% |  | (22) |
| 65+ | 5.5% |  | (22) |
| HIV risk behaviors | **Female** | **Male** |  |
| Abstinent, past year | 49.9% | 22.9% | (23) |
| Low risk (monogamous relationships) | 40.4% | 61.5% | (23-25) |
| Moderate risk (2-3 concurrent relationship) | 5.1% | 11.8% | (23-25) |
| High risk (>3 concurrent relationships) | 4.6% | 3.8% | (26-28) |
| **HIV transmission and risk behaviors** |  |  |  |
| Annual diagnosis probability (if undiagnosed) | 67.0% |  | (29) |
| Annual probability of linkage to care after a positive HIV test |  |  |  |
| Chronic stage (CD4 ≥ 200) | 88.9% |  | (30, 31) |
| Advanced stage (CD4 < 200) | 59.2% |  | (30, 31) |
| Probability of adherence to ART (proportion of doses taken as directed) | 63.0% |  | (32-36) |
| Annual probability of ART regimen change in response to resistance | 58.1% |  | (35) |
| Probability of virologic suppression |  |  |  |
| Perfect adherence | 95% |  | (37) |
| Average adherence | 60% |  | (32-37) |
| Probability of condom use, combined across settings and partners | 60% |  | [S15] |
| Sexual risk behaviors |  |  |  |
| Annual rate of partner change | **Female** | **Male** |  |
| Low risk | 0.084 | 0.296 | (38, 39) |
| Moderate risk | 0.350 | 0.337 | (39) |
| High risk | 1.748 | 0.591 | (38, 39) |
| Assortativity of sexual mixing by |  |  |  |
| Partner change rate | 0.32 |  | (40) |
| Age | 0.29 |  | (40) |
| CASM | 0.43 |  | (40) |
| **Programmatic cost per intervention per person per year (US \$)** | | | |
| VMMC | \$ 132.90 | | (41) |
| Long-Acting PrEP | \$ 144.00 | | (42) |
| Oral PrEP | \$ 87.52 | | [Administrative Data] |
| Condom distribution | \$ 6.74 | | (41) |
| DREAMS | \$ 29.16 | | [Administrative Data] |
| Community-based testing and counseling | \$ 33.00 | | (43) |

|  |  |  |
| --- | --- | --- |
| Index tracing | \$ 24.27 | [Administrative Data] |
| HIV Self-Testing (per test) | \$ 5.70 | (44) |
| Defaulter tracing | \$ 25.16 | [Administrative Data] |
| Differentiated care | \$ 19.47 | (45), [Calculated] |
| ART adherence support | \$ 31.61 | [Calculated] |

**Table 2.** Changes in model inputs with discontinuation of USAID and CDC funding.

| HIV outcomes impacted by interventions | Current Coverage | Coverage after funding cuts |  |  |
| --- | --- | --- | --- | --- |
|  |  | - USAID funds + CDC funds | + USAID funds - CDC funds | - USAID funds - CDC funds |
| ART coverage | 86% | 84.3% | 80.0% | 78.3% |
| Voluntary Medical Male Circumcision (VMMC) – 1 time | 35% | 1.5% | 0.3% | 0.3% |
| Oral PrEP | 13% | 12.5% | 12.6% | 12.1% |
| Condom distribution | 69% | 61.7% | 61.7% | 54.3% |
| Community-based testing and counseling | 18% | 15.8% | 14.8% | 12.6% |
| Index tracing | 43.0% | 37.4% | 37.4% | 31.8% |
| HIV self-testing | 1% | 0.7% | 0.5% | 0.2% |
| Defaulter tracing | 48% | 41.5% | 43.2% | 36.7% |
| Differentiated care | 50% | 43.3% | 45.0% | 38.2% |
| ART adherence support | 60% | 51.9% | 53.9% | 45.9% |

Outcomes of interest included increases in the HIV incidence rate, the number of new infections, and mortality from those new infections. We used a compartmental transmission model that simulated the effects of HIV interventions on transmission, diagnosis, and viral load suppression.(12) Compartments included sex (male/female), age in years (<15/15-24/25-34/35-64/≥65), HIV-risk types for men (men who have sex with men/non-MSM), HIV-risk levels (low risk/medium risk/high risk), genotypic resistance (wildtype HIV/resistant HIV), and presence vs. absence of CASM (Conditions of Alcohol, Substance and Mood disorders) conditions. High risk groups were defined as people with large numbers of concurrent sexual partners (e.g., transactional sex workers), medium risk groups were others who were in non-monogamous relationships in the past year, and low risk groups were sexually active people in monogamous relationships. Risk levels, CASM status, and risk types for men (i.e., MSM, non-MSM) impact transmissibility through differing frequencies and/or risks of sexual contacts.

Vertical transmission and horizontal sexual transmission are only considered, as injection drug use is uncommon in KZN. Our model mirrors the course of HIV infection through five sequential stages: susceptible, acute HIV, chronic HIV, advanced HIV, and HIV-related death. Each of the HIV stages is further subdivided by diagnosis status (“yes” versus “no”), ART treatment status (“yes” versus “no,” and, if “yes,” further modified by adherence and retention-in-care), and viral load suppression (“yes” versus “no”). Scaling up a particular HIV intervention produces corresponding changes in how the population distributes among model compartments representing diagnosis status, treatment status, and/or viral load suppression, impacting transmissibility, morbidity, and mortality.

We explored various scenarios for restoring funding at 12 months and paused through 2030, extent (USAID-implemented, CDC-implemented, or both), and potential for reallocation (i.e., oral PrEP funding reallocated to the more cost-effective long-acting PrEP).

Intervention costs were assessed using an ingredients-based micro-costing approach, encompassing materials (e.g. medication doses) and infrastructure (e.g., stocking and delivery-related expenditures). We made simplifying assumptions of instantaneous funding cessations and restorations, ignoring stockpiles of equipment and frictional costs for deactivating and activating labor. We also made the simplifying assumption that treatment effectiveness exhibited constant decreasing returns to scale (e.g. 80% funding produced 80% of the effect). Model inputs regarding baseline intervention coverage and effectiveness on HIV incidence, detection, treatment initiation, and treatment success were adapted to KZN based on prior published work.(12) For example, in “real world” settings, oral PrEP prevents relatively few HIV infections during the year after initiation because daily adherence is low and retention is 10%-30%,(13-17) whereas long-acting PrEP prevents more HIV infections during the year after initiation because daily adherence is not required and retention is 60%-90%.(18)

## RESULTS

In KZN province, South Africa, we estimated the 12-month, and through 2030 USAID and CDC funding cuts (Figure) would increase HIV incidence by 3.4%, and 22.8%, respectively, by 2030; thus, contributing to approximately 35,300 and 116,100 additional infections, respectively, through 2030; and result in approximately 12,800 and 42,300 additional deaths, respectively. Approximately 31% of added infections would be attributable to decreases in ART supply, and approximately 43% of added infections would be attributable to USAID cuts alone (Figure B). We estimate USAID and CDC funding cuts in KZN totaling $59.3M per year (Table 2). Of this $59.3M, USAID contributes approximately $22.8M per year and CDC contributes approximately $36.5M per year.

**Figure.**
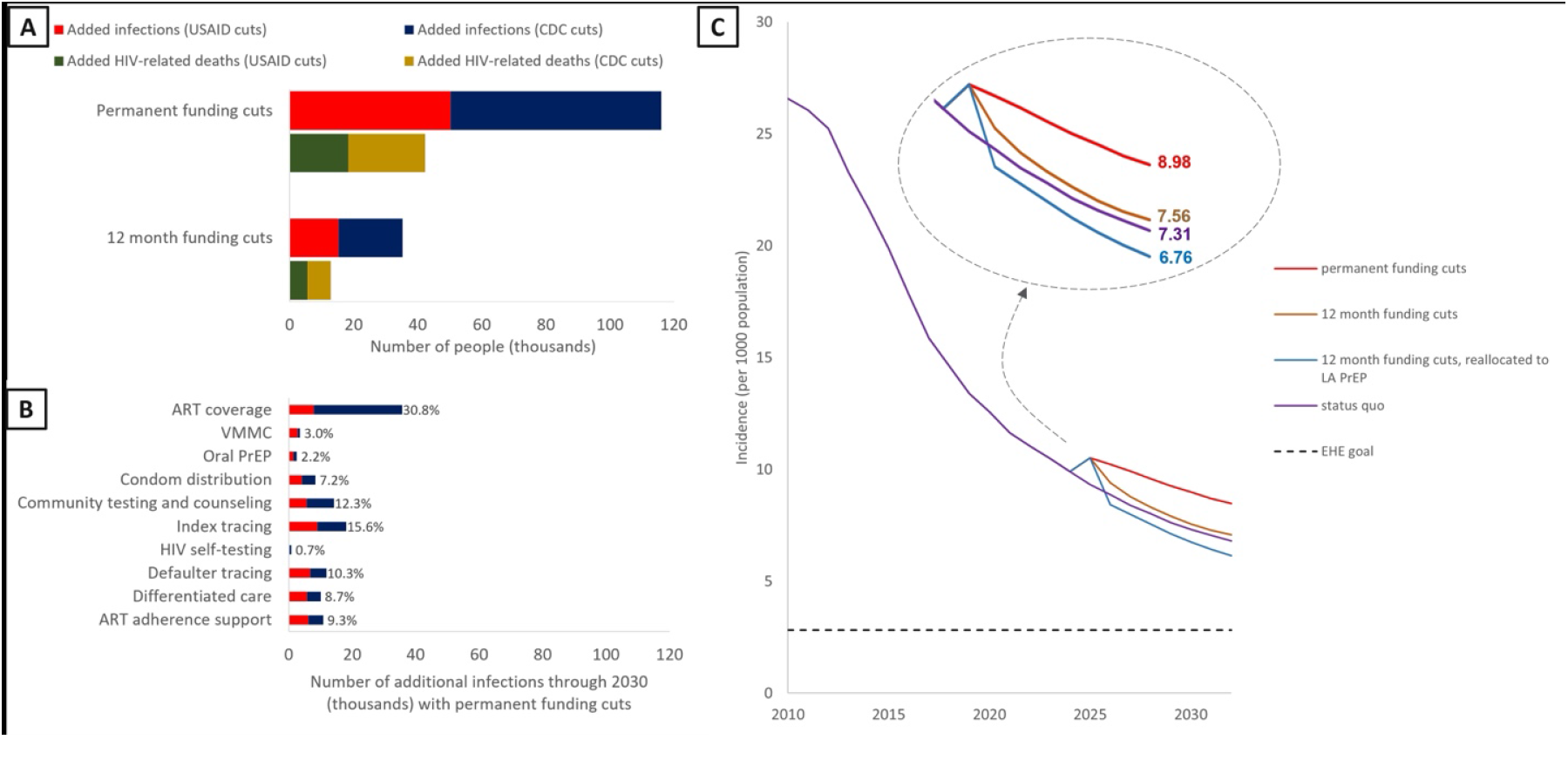
Projected impact of USAID and CDC funding cuts on HIV incidence and mortality. A) Model estimations of additional HIV infections and HIV-related deaths through 2030, plotted in 12-month interval, resulting from USAID and CDC funding cuts across various interventions for specified durations of 12 months and through 2030. The model applied the coverage reductions in the Table for the specified durations, after which coverage was assumed to be instantaneously and completely restored. USAID funding cuts are distinguished from CDC funding cuts. B) Model estimations of additional HIV infections resulting from permanent USAID and CDC funding cuts, stratified across interventions. C) Model estimations of changes in HIV incidence resulting from USAID and CDC funding reductions across various interventions for specified durations of 12 months and by 2030. Restoration was evaluated without reallocation of funds, and with reallocation of funds from oral PrEP to long-acting PrEP (LA PrEP).

If the refunding process permits reallocation of resources, some of these additional infections could be averted through gains in efficiency (Figure C). Reallocating funds for oral PrEP towards long-acting PrEP could potentially offset the increase in HIV incidence that would occur by 2030 from funding pauses of 12 months or less, assuming that the annual supply and dispensing costs for long-acting PrEP were no greater than 1.5 times of those of oral PrEP. After a 12-month pause (Figure), reallocation of funds toward long-acting PrEP could avert 12,600 additional infections.

## DISCUSSION

Withholding CDC and USAID funding threatens the tremendous progress in addressing the HIV epidemic in South Africa and particularly KZN province. HIV incidence will increase by more than 3% within a year and by nearly 23% by 2030, and more than 40,000 additional deaths will occur by 2030. Restoring CDC and USAID funding within 12 months would mitigate the threat, and if combined with more efficient reallocation, and the infrastructure to support it, could offset the threat and permit KZN to progress again toward achieving the Ending the HIV Epidemic (EHE) goals.

We found less proportional impact of USAID and CDC funding cuts on HIV infections than Gandhi et al’s modeling study in South Africa (23% versus 50%), but that study did not assess programmatic impact on funding of particular interventions.(19) We also found lesser impact than the range (100% - 350%) predicted for sub-Saharan African countries in aggregate by a global HIV modeling platform (OPTIMA).(20) Our estimate was consistent with best-case scenario results from a South African modeling study (29% increase in HIV infections over 4 years) that assumed reductions in program funding would be proportionate to the percentage funded by PEPFAR in 2023.(21) None of these studies evaluated scenarios in which funding was quickly restored.

Limitations of this analysis include simplifications from assuming instantaneous intervention scale-down and scale-up, constant returns to scale, and stability of Global Fund donations to KZN, even though diminishing U.S. contributions may reduce future donations. As a rapid assessment, exhaustive sensitivity analyses were not conducted. Additionally, we did not incorporate operational delays, workforce losses, infrastructure modifications, or supply chain reactivation challenges that would be associated with funding cuts and potential reallocation of funding. However, these limitations are offset by important strengths, including considering observed changes in programmatic coverage rather than assuming hypothetical across-the-board USAID and CDC cuts, considered offsets from task-shifting and reallocations of domestic resources, using a KZN-specific model, and examining alternative restoration scenarios regarding timing, extent, and reallocation of resources.

## CONCLUSION

Withdrawal of USAID and CDC commitments in the largest HIV epidemic in the world leads to increased incidence and mortality and threatens not only KZN, South Africa but global HIV control. Our study indicates that restoring funding within 12 months and increasing efficiency of HIV interventions can reestablish KZN South Africa’s trajectory toward EHE goals.

## Data Availability

All scientific data used to populate the simulation model are derived from existing, secondary data, including published scientific articles, nationally representative country-level reports, and Ministry of Health and HIV service provider sources and listed in the mansucript. Data are publicly available, cited, or are available upon request. Modeling outcomes will be shared via peer-review publication with results from additional modeling analyses available upon request. Combination C++ and Python modeling code will be stored on NYU servers and will be available upon request.

## References

1. Ndlovu N, Mokganya M, Blose N, Padarath A, editors.. District Health Barometer 2023/2024. Durban: Health Systems Trust; 2025.

2. Shisana O, Rehle, T, Simbayi LC, Parker W, Zuma, K, Bhana A, Connolley C, Jooste S, PIllay V et al. South African National Prevalence, Incidence, Behavior, and Communication Survey. Cape Town: HSRC Press; 2005.

3. Zuma K, Zungu NP, Moyo S, Marinda E, Simbayi LC, Jooste SE, Mabaso M, Ramlagan S, Makola L, van Zyl J, Naidoo I and the SABSSMVI team.. The Sixth South African National HIV, e Sixth South African National HIV, Behavioural and Health Survey 2022: A Summary Report. Cape Town HSRC Press 2024.

4. Institute fSS. South Africa country profile. African Futures and Innovation. [Available from: https://futures.issafrica.org/geographic/countries/southafrica.

5. Abrams MP, Weiner J, Piske M, Enns B, Krebs E, Zang X, et al. Translating and disseminating a localised economic model to support implementation of the ‘Ending the HIV Epidemic’ initiative to public health policymakers. Evid Policy. 2023;19(4):554–71.

6. Basu S, Yudkin JS, Jawad M, Ghattas H, Hamad BA, Jamaluddine Z, et al. Reducing non-communicable diseases among Palestinian populations in Gaza: A participatory comparative and cost-effectiveness modeling assessment. PLOS Glob Public Health. 2024;4(5):e0003168.

7. Bellizzi S, Letchford N, Adib K, Probert WJM, Hancock P, Alsawalha L, et al. Participatory Mathematical Modeling Approach for Policymaking during the First Year of the COVID-19 Crisis, Jordan. Emerg Infect Dis. 2023;29(9):1738–46.

8. Holtgrave DR, Maulsby C, Kim JJ, Cassidy-Stewart H, Hauck H. Using Resource Allocation Modeling to Inform HIV Prevention Priority Setting for Baltimore-Towson, Maryland. Prog Community Health Partnersh. 2016;10(1):133–9.

9. Ngcamphalala C, Yeo YR, Su JI, Jeetoo M, Charles D, Tfwala Z, Mahlalela N, Dube L, Sahabo R, Dlamini T, Nuwagaba-Biribonwoha H, Braithwaite RS.. Determining minimum resources required by Eswatini to meet its Ending the HIV Epidemic 2030 goal. JIAS.in press.

10. Apollo T, Yeo YR, Mugurungi OM, Sithole N, Taramusi I, Tachiwenyika E, Gwavava EP, Makoni W, Chimwaza A, Takarinda KC, Ncube G, Jeetoo M, Charles D, Braithwaite RS. Case study of PEPFAR funding cuts on HIV infections and deaths: reversing hard-won gains. AIDS. 2025;39(14):2143–6.

11. Jeetoo M, Charles D, Yeo YR, Su J, Mbaya JK, Shenoi S, Morojele N, Moll T, Braithwaite RS.. Addressing alcohol use and depression as a strategy to reduce HIV in KwaZulu-Natal, South Africa. Conference on Retroviruses and Opportunistic Infections (CROI) 2025; San Francisco, CA, 2025.

12. Su JI, Yeo YR, Jeetoo M, Morojele NK, Francis JM, Shenoi S, et al. Cost-effectiveness of screening and treating alcohol use and depression among people living with HIV in Zimbabwe: a mathematical modeling study. BMC Med. 2024;22(1):481.

13. de Dieu Tapsoba J, Zangeneh SZ, Appelmans E, Pasalar S, Mori K, Peng L, et al. Persistence of oral pre-exposure prophylaxis (PrEP) among adolescent girls and young women initiating PrEP for HIV prevention in Kenya. AIDS Care. 2021;33(6):712–20.

14. Hendrickson C, Hirasen K, Mongwenyana C, Benade M, Bothma R, Smith C, et al. Costs and outcomes of routine HIV oral pre-exposure prophylaxis implementation across different service delivery models and key populations in South Africa: a retrospective cohort study. The lancet HIV. 2025;12(2):e130–e42.

15. Kagaayi J, Batte J, Nakawooya H, Kigozi B, Nakigozi G, Strömdahl S, et al. Uptake and retention on HIV pre-exposure prophylaxis among key and priority populations in South-Central Uganda. Journal of the International AIDS Society. 2020;23(8):e25588.

16. Mayanja Y, Kamacooko O, Lunkuse JF, Muturi-Kioi V, Buzibye A, Omali D, et al. Oral pre-exposure prophylaxis preference, uptake, adherence and continuation among adolescent girls and young women in Kampala, Uganda: a prospective cohort study. Journal of the International AIDS Society. 2022;25(5):e25909.

17. Rousseau E, Wu L, Heffron R, Baeten JM, Celum CL, Travill D, et al. Association of sexual relationship power with PrEP persistence and other sexual health outcomes among adolescent and young women in Kenya and South Africa. Front Reprod Health. 2023;5:1073103.

18. Spinelli MA, Bisom-Rapp E, Heise MJ, Camp C, Appa A, Liu AY, et al. High Retention and Adherence With Rapid Long-acting Injectable Preexposure Prophylaxis Implementation in an Urban Safety Net Clinic Population. Clin Infect Dis. 2025;80(5):1139–42.

19. Gandhi AR, Bekker LG, Paltiel AD, Hyle EP, Ciaranello AL, Pillay Y, et al. Potential Clinical and Economic Impacts of Cutbacks in the President’s Emergency Plan for AIDS Relief Program in South Africa : A Modeling Analysis. Ann Intern Med. 2025;178(4):457–67.

20. Brink DT, Martin-Hughes R, Bowring AL, Wulan N, Burke K, Tidhar T, et al. Impact of an international HIV funding crisis on HIV infections and mortality in low-income and middle-income countries: a modelling study. The lancet HIV. 2025;12(5):e346–e54.

21. Stover J, Sonneveldt E, Tam Y, Horton KC, Phillips AN, Smith J, et al. Effects of reductions in US foreign assistance on HIV, tuberculosis, family planning, and maternal and child health: a modelling study. The Lancet Global Health. 2025;13(10):e1669–e80.

22. Statistical Release Mid-year Population Estimates. South Africa: Statistics South Africa, Republic of South Africa 2021.

23. Herbst DK, Pillay PD, Baisley MK, Barnighausen PT, Dlamini MS, Harling DG, et al. South Africa - DREAMS baseline study in females aged 15-24 years in rural KwaZulu Natal, South Africa, 2006–2015. In: Herbst DK, Pillay PD, Baisley MK, Barnighausen PT, Dlamini MS, Harling DG, et al., editors. v1.0.0 ed. South Africa2006-2015.

24. South Africa Demorgaphic and Health Survey 2016 National Department of Health (NDoH), Statistics South Africa (Stats SA), South African Medical Research Council (SAMRC), and ICF.

25. Herbst DK, Pillay PD, Baisley MK, Barnighausen PT, Dlamini MS, Harling DG, et al. South Africa - DREAMS baseline study in young men aged 20-29 years in rural KwaZulu Natal, South Africa, 2006–2015. In: Herbst DK, Pillay PD, Baisley MK, Barnighausen PT, Dlamini MS, Harling DG, et al., editors. South Africa2006-2015.

26. Chimbindi NZ, McGrath N, Herbst K, San Tint K, Newell ML. Socio-Demographic Determinants of Condom Use Among Sexually Active Young Adults in Rural KwaZulu-Natal, South Africa. Open AIDS J. 2010;4:88–95.

27. Bello B, Moultrie H, Somji A, Chersich MF, Watts C, Delany-Moretlwe S. Alcohol use and sexual risk behaviour among men and women in inner-city Johannesburg, South Africa. BMC Public Health. 2017;17(Suppl 3):548.

28. Patterns of Men’s Reproductive and Sexual Behaviour in South Africa. Statistics South Africa Republic of South Africa. Report No.: 03-10-21.

29. Simbayi LC, Zuma K, Zungu N, Moyo S, Marinda E, Jooste S, Mabaso M,, Ramlagan S NA, van Zyl J, Mohlabane N, Dietrich C, Naidoo I and the SABSSM V Team. South African National HIV Prevalence, Incidence, Behaviour and Communication Survey, 2017. Cape Town, South Africa; 2019.

30. Perelman J, Rosado R, Ferro A, Aguiar P. Linkage to HIV care and its determinants in the late HAART era: a systematic review and meta-analysis. AIDS Care. 2018;30(6):672–87.

31. Zetola NM, Bernstein K, Ahrens K, Marcus JL, Philip S, Nieri G, et al. Using surveillance data to monitor entry into care of newly diagnosed HIV-infected persons: San Francisco, 2006-2007. BMC Public Health. 2009;9:17.

32. Sithole Z, Mbizvo E, Chonzi P, Mungati M, Juru TP, Shambira G, et al. Virological failure among adolescents on ART, Harare City, 2017-a case-control study. BMC Infect Dis. 2018;18(1):469.

33. Erlwanger AS, Joseph J, Gotora T, Muzunze B, Orne-Gliemann J, Mukungunugwa S, et al. Patterns of HIV Care Clinic Attendance and Adherence to Antiretroviral Therapy Among Pregnant and Breastfeeding Women Living With HIV in the Context of Option B+ in Zimbabwe. J Acquir Immune Defic Syndr. 2017;75 Suppl 2:S198–s206.

34. Vreeman RC, Scanlon ML, Tu W, Slaven JE, McAteer CI, Kerr SJ, et al. Validation of a self-report adherence measurement tool among a multinational cohort of children living with HIV in Kenya, South Africa and Thailand. J Int AIDS Soc. 2019;22(5):e25304.

35. Haas AD, Msukwa MT, Egger M, Tenthani L, Tweya H, Jahn A, et al. Adherence to Antiretroviral Therapy During and After Pregnancy: Cohort Study on Women Receiving Care in Malawi’s Option B+ Program. Clin Infect Dis. 2016;63(9):1227–35.

36. Davies MA, Boulle A, Fakir T, Nuttall J, Eley B. Adherence to antiretroviral therapy in young children in Cape Town, South Africa, measured by medication return and caregiver self-report: a prospective cohort study. BMC Pediatr. 2008;8:34.

37. Byrd KK, Hou JG, Hazen R, Kirkham H, Suzuki S, Clay PG, et al. Antiretroviral Adherence Level Necessary for HIV Viral Suppression Using Real-World Data. J Acquir Immune Defic Syndr. 2019;82(3):245–51.

38. Nsubuga RN, White RG, Mayanja BN, Shafer LA. Estimation of the HIV basic reproduction number in rural south west Uganda: 1991-2008. PLoS One. 2014;9(1):e83778.

39. Harrison A, Cleland J, Frohlich J. Young people’s sexual partnerships in KwaZulu-Natal, South Africa: patterns, contextual influences, and HIV risk. Stud Fam Plann. 2008;39(4):295–308.

40. Malagón T, Burchell A, El-Zein M, Tellier PP, Coutlée F, Franco EL. Assortativity and Mixing by Sexual Behaviors and Sociodemographic Characteristics in Young Adult Heterosexual Dating Partnerships. Sex Transm Dis. 2017;44(6):329–37.

41. Meyer-Rath G, van Rensburg C, Chiu C, Leuner R, Jamieson L, Cohen S. The per-patient costs of HIV services in South Africa: Systematic review and application in the South African HIV Investment Case. PLoS One. 2019;14(2):e0210497.

42. Smith J, Bansi-Matharu L, Cambiano V, Dimitrov D, Bershteyn A, van de Vijver D, et al. Predicted effects of the introduction of long-acting injectable cabotegravir pre-exposure prophylaxis in sub-Saharan Africa: a modelling study. Lancet HIV. 2023;10(4):e254–e65.

43. Minnery M, Mathabela N, Shubber Z, Mabuza K, Gorgens M, Cheikh N, et al. Opportunities for improved HIV prevention and treatment through budget optimization in Eswatini. PLoS One. 2020;15(7):e0235664.

44. Jamieson L, Johnson LF, Matsimela K, Sande LA, d’Elbée M, Majam M, et al. The cost effectiveness and optimal configuration of HIV self-test distribution in South Africa: a model analysis. BMJ Glob Health. 2021;6(Suppl 4).

45. Rosen S, Nichols B, Guthrie T, Benade M, Kuchukhidze S, Long L. Do differentiated service delivery models for HIV treatment in sub-Saharan Africa save money? Synthesis of evidence from field studies conducted in sub-Saharan Africa in 2017-2019. Gates Open Res. 2021;5:177.

